# Report 20: Using mobility to estimate the transmission intensity of COVID-19 in Italy: A subnational analysis with future scenarios

**DOI:** 10.1101/2020.05.05.20089359

**Authors:** Michaela A. C. Vollmer, Swapnil Mishra, H Juliette T Unwin, Axel Gandy, Thomas A. Mellan, Valerie Bradley, Harrison Zhu, Helen Coupland, Iwona Hawryluk, Michael Hutchinson, Oliver Ratmann, Melodie Monod, Patrick Walker, Charlie Whittaker, Lorenzo Cattarino, Constance Ciavarella, Lucia Cilloni, Kylie Ainslie, Marc Baguelin, Sangeeta Bhatia, Adhiratha Boonyasiri, Nicholas Brazeau, Giovanni Charles, Laura V Cooper, Zulma Cucunuba, Gina Cuomo-Dannenburg, Amy Dighe, Bimandra Djaafara, Jeff Eaton, Sabine L van Elsland, Richard FitzJohn, Keith Fraser, Katy Gaythorpe, Will Green, Sarah Hayes, Natsuko Imai, Ben Jeffrey, Edward Knock, Daniel Laydon, John Lees, Tara Mangal, Andria Mousa, Gemma Nedjati-Gilani, Pierre Nouvellet, Daniela Olivera, Kris V Parag, Michael Pickles, Hayley A Thompson, Robert Verity, Caroline Walters, Haowei Wang, Yuanrong Wang, Oliver J Watson, Lilith Whittles, Xiaoyue Xi, Azra Ghani, Steven M Riley, Lucy Okell, Christl A. Donnelly, Neil M Ferguson, Ilaria Dorigatti, Seth Flaxman, Samir Bhatt

**Author notes:** Contributed equally (Michaela A. C. Vollmer, Swapnil Mishra, H Juliette T Unwin, Axel Gandy, Seth Flaxman, Samir Bhatt).

## Abstract

Italy was the first European country to experience sustained local transmission of COVID-19. As of 1st May 2020, the Italian health authorities reported 28,238 deaths nationally. To control the epidemic, the Italian government implemented a suite of non-pharmaceutical interventions (NPIs), including school and university closures, social distancing and full lockdown involving banning of public gatherings and non essential movement. In this report, we model the effect of NPIs on transmission using data on average mobility. We estimate that the average reproduction number (a measure of transmission intensity) is currently below one for all Italian regions, and significantly so for the majority of the regions. Despite the large number of deaths, the proportion of population that has been infected by SARS-CoV-2 (the attack rate) is far from the herd immunity threshold in all Italian regions, with the highest attack rate observed in Lombardy (13.18% [10.66%-16.70%]). Italy is set to relax the currently implemented NPIs from 4th May 2020. Given the control achieved by NPIs, we consider three scenarios for the next 8 weeks: a scenario in which mobility remains the same as during the lockdown, a scenario in which mobility returns to pre-lockdown levels by 20%, and a scenario in which mobility returns to pre-lockdown levels by 40%. The scenarios explored assume that mobility is scaled evenly across all dimensions, that behaviour stays the same as before NPIs were implemented, that no pharmaceutical interventions are introduced, and it does not include transmission reduction from contact tracing, testing and the isolation of confirmed or suspected cases. New interventions, such as enhanced testing and contact tracing are going to be introduced and will likely contribute to reductions in transmission; therefore our estimates should be viewed as pessimistic projections. We find that, in the absence of additional interventions, even a 20% return to pre-lockdown mobility could lead to a resurgence in the number of deaths far greater than experienced in the current wave in several regions. Future increases in the number of deaths will lag behind the increase in transmission intensity and so a second wave will not be immediately apparent from just monitoring of the daily number of deaths. Our results suggest that SARS-CoV-2 transmission as well as mobility should be closely monitored in the next weeks and months. To compensate for the increase in mobility that will occur due to the relaxation of the currently implemented NPIs, adherence to the recommended social distancing measures alongside enhanced community surveillance including swab testing, contact tracing and the early isolation of infections are of paramount importance to reduce the risk of resurgence in transmission.

**SUGGESTED CITATION:** Michaela A. C. Vollmer, Swapnil Mishra, H Juliette T Unwin, Axel Gandy *et al*. Using mobility to estimate the transmission intensity of COVID-19 in Italy: a subnational analysis with future scenarios. Imperial College London (2020) doi:https://doi.org/10.25561/78677

This work is licensed under a Creative Commons Attribution-NonCommercial-NoDerivatives 4.0 International License.

## 1 Introduction

Following the emergence of a novel coronavirus (SARS-CoV-2) and its spread outside of China, Italy was the first European country to be hit by COVID-19. As of 1*st* May 2020, 28,238 deaths have been reported nationally with 13,860 having occurred in Lombardy, the most populous and worst hit region in Italy. In Lombardy, infection has been estimated to be introduced in early January and transmission went undetected until the first confirmed case of COVID-19 was reported on 20th of February [3]. On the 21st of February case testing began in order to trace new SARS-CoV-2 infections, and the first COVID-19 death was reported on the 23rd of February in Vo, in the Veneto region [11].

In response to a rapid escalation of hospital demand and deaths, unprecedented non-pharmaceutical interventions (NPIs) were implemented first in Lombardy, Veneto and some neighbouring regions [3] and then extended to all of Italy. The goal of these interventions was to control the epidemic, reduce healthcare demand, and minimise pressure on the national health system. The interventions implemented in Italy included case isolation, the closure of schools and universities, banning of mass gatherings and public events, ban of movement and wide-scale social distancing.

Mathematical and statistical models are useful tools to better understand the transmission dynamics of infectious diseases. They can assess the dynamics of an epidemic as it evolves in time, evaluate the impact of interventions and simulate future scenarios. While models often rely on noisy epidemiological data, they can be designed to account for the uncertainties in the data and represent conceptual frameworks that can be used to look at trends, infer dynamics and answer real-world questions using an evidence-based approach. The real-time analysis and modelling of epidemic data can thus provide data-driven and scientific evidence that can inform the response, planning and public health decision making against the current COVID-19 pandemic.

The recent release of mobility data by Google [1] is a useful resource to measure the impact of the interventions implemented against COVID-19. These data provide fine-grained, population-wide information on the relative changes in movement, and can be used to measure the transmissibility of SARS-CoV-2 by acting as a proxy for changes in behaviour. The Google data we use have been collected by geographical location in categories of retail and recreation, groceries and pharmacies, parks, transit stations, workplaces and residential.

In this report we analyse the incidence of death reported across the 20 Italian regions, and along with the observed relative changes in regional movement, assess how interventions have impacted the transmissibility of SARS-CoV-2. We provide estimates of the number of deaths averted by the implementation of the control measures, the expected proportion of population infected (as of 1*st* May 2020), and explore the potential impact that the relaxation of the current interventions could have on disease transmission in the future. Understanding what impact the relaxation of the currently implemented NPIs (‘exit strategies’) will have on transmission is critical in guiding policy decisions to manage the transmission of COVID-19 in the so-called ‘Phase 2’.

**Figure 1:**
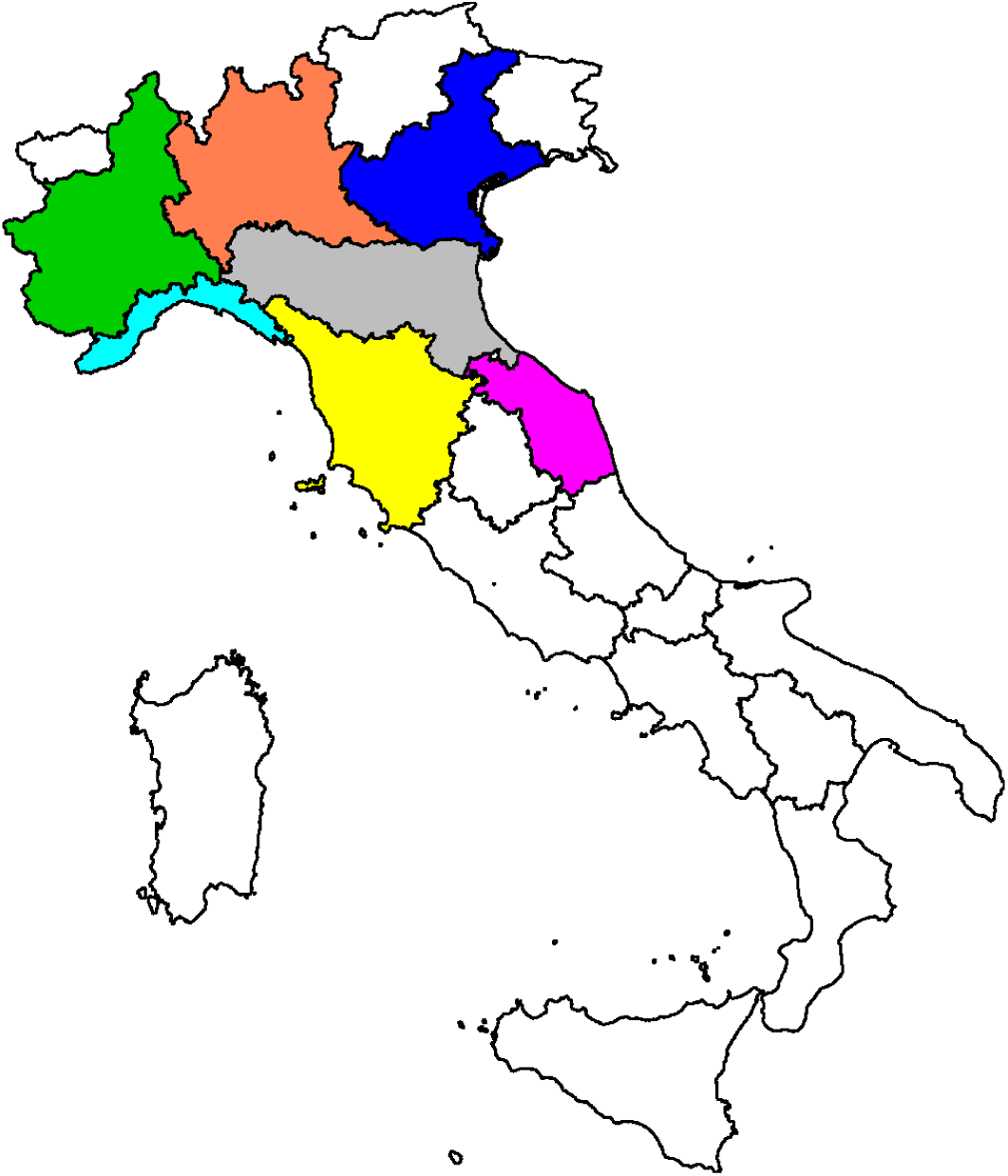
The seven highest COVID-19 mortality areas of Italy (in terms of the absolute number of deaths): Lombardy (Orange), Emilia-Romagna (Gray), Piedmont (Green), Veneto (Blue), Liguria (Cyan), Marche (Pink) and Tuscany (Yellow).

## 2 Results

### 2.1 Using mobility to inform transmission

Figure 2 shows trends in mobility from Google data at a regional level for the 7 regions with more than 500 COVID-19 deaths (see Table 1), which are Lombardy, Emilia-Romagna, Piedmont, Veneto, Liguria, Marche and Tuscany. The mobility dimensions are detailed in Section 4. Briefly, the mobility data show the relative change in mobility with respect to a baseline calculated shortly before the COVID-19 epidemic within each region. A value of say –0.2 in the retail and recreation section means that individuals, on average, are visiting retail and recreation locations 20% less than before the epidemic. In Figure 2 we also overlay the timing of major NPIs (Appendix Table 6.3). Due to very strong collinearity across mobility dimensions we only use residential, transit stations, and an average of the remaining four dimensions (i.e. retail and recreation, groceries and pharmacies, parks, and workplaces). The residential dimension is a proxy for household transmission and the transit dimension is a proxy for general travel within and between regions, including time spent at travel hubs. The average mobility is the mean of the other dimensions and is a proxy for general day-to-day activities. There is clear visual correspondence between the dates interventions were implemented and the observed reductions in mobility. This is demonstrated statistically by the large mean correlation of 0.81 obtained with a simple linear model regressing interventions (as piecewise constant) on the average mobility dimension. This suggests that mobility can act as a suitable proxy for the changes in behaviour induced by the implementation of the major NPIs. We do note however, that mobility does not capture all the heterogeneity in transmission, specifically missing factors such as case-based interventions and the effect of school and university closures.

**Table 1:**
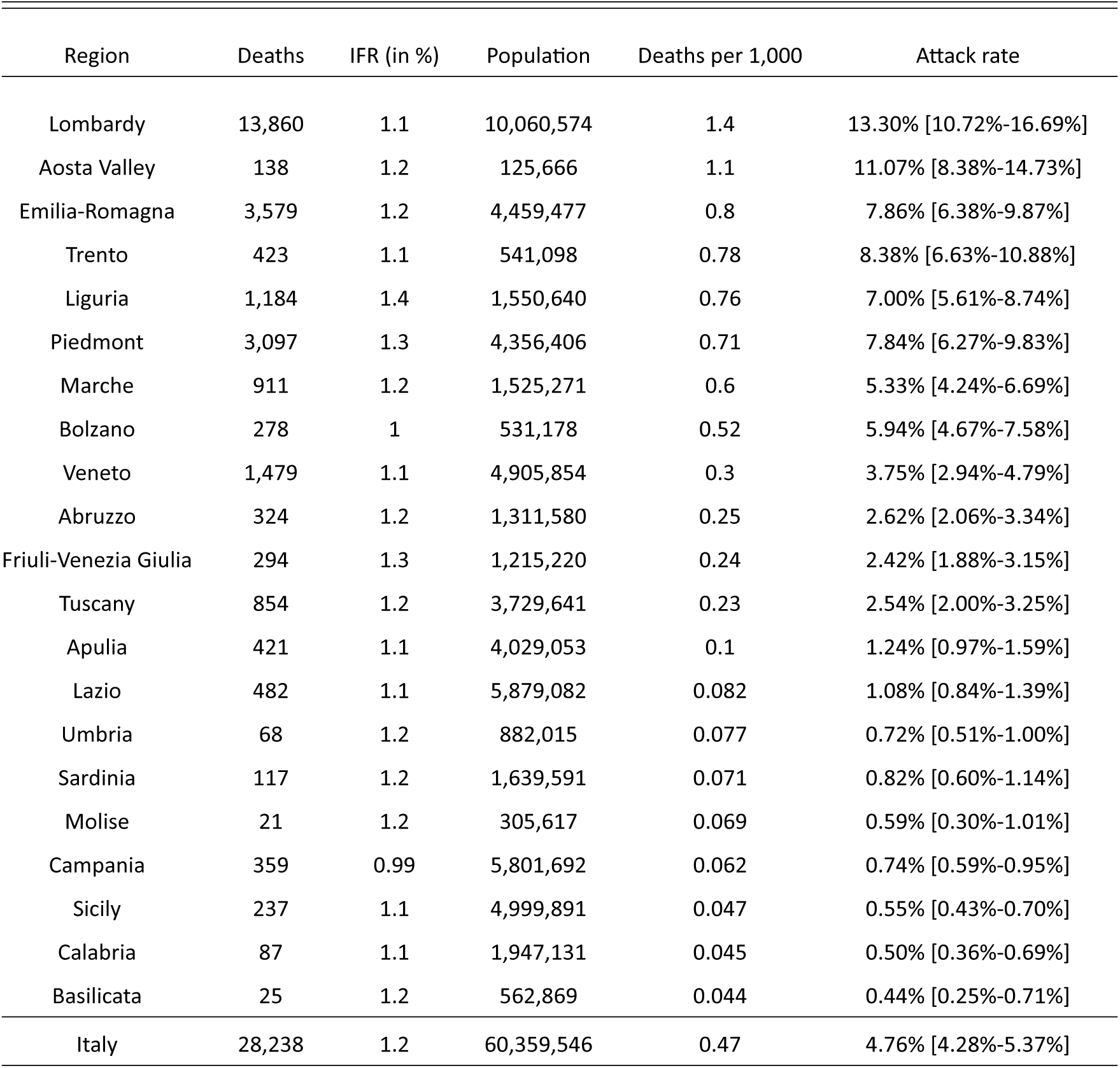
Table listing reported deaths, estimated infection fatality rate (*IFR*), population, deaths per capita, and our estimates of the attack rate (percent of the population infected) for all regions in Italy

**Figure 2:**
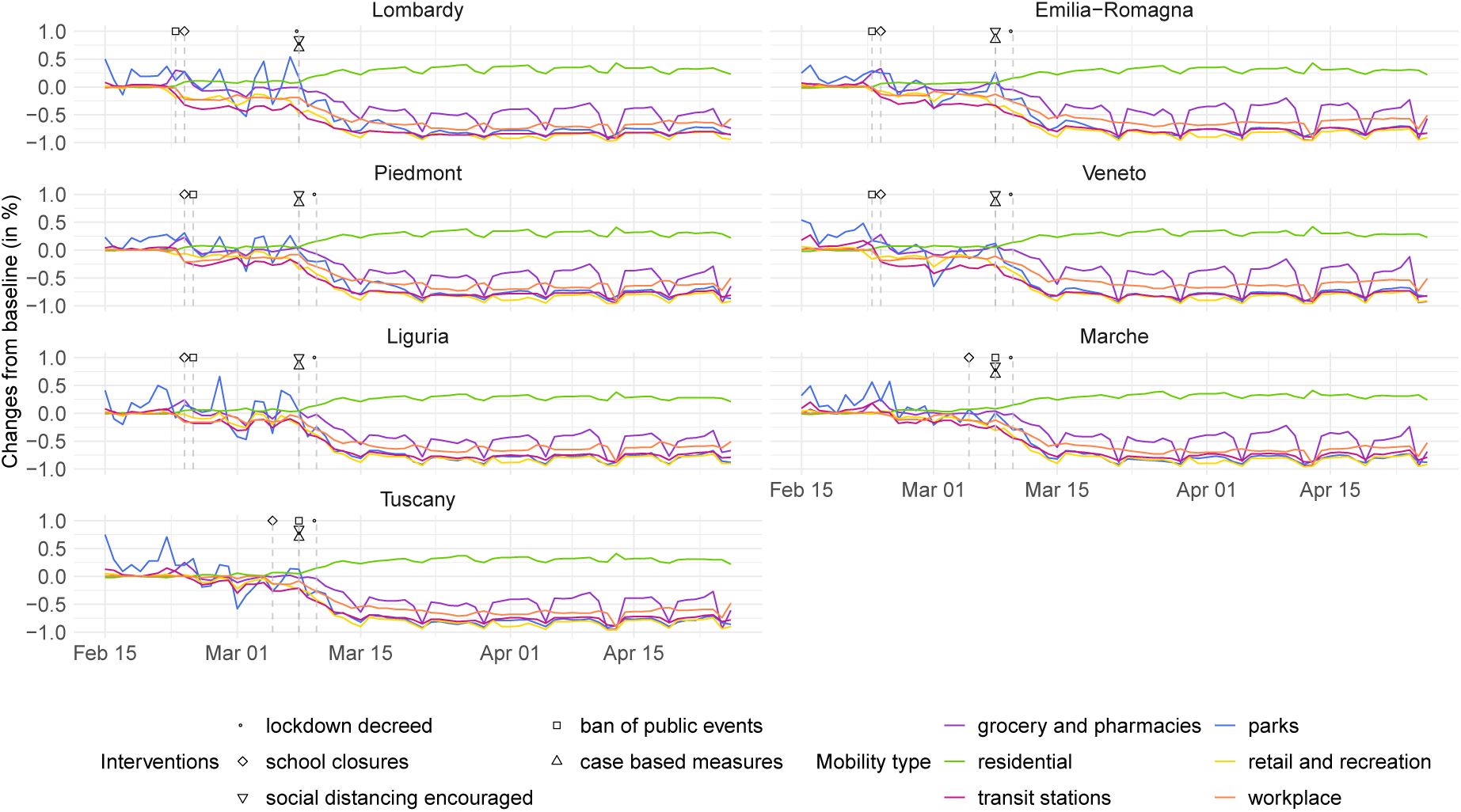
Interventions and mobility data for the seven most-affected regions in Italy.

Figure 3 shows the average global effect sizes for the mobility dimensions used in our model. Due to collinearity, it is not statistically possible to identify which dimension has had the largest impact on *R_t_*. However, we do find that the transit dimension and the average mobility dimension are statistically significant, while the residential dimension is not (though the posterior mean is less than 0). We hypothesise that the residential covariate could increase *R_t_* due to household transmission between cohabitants.

**Figure 3:**
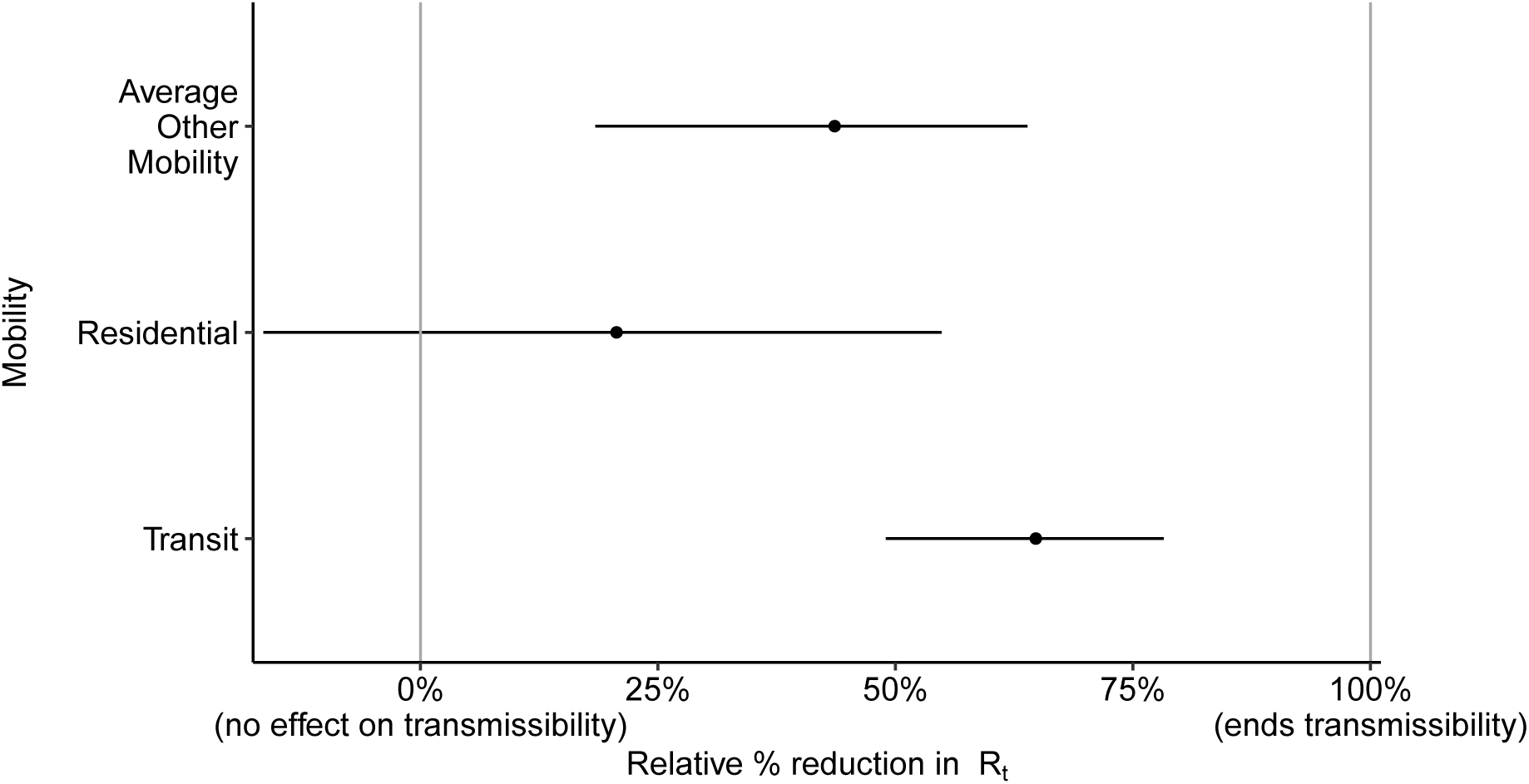
Mobility effect sizes: relative reduction in *R_t_* if the specified mobility was completely stopped.

Figure 9 in the Appendix shows the partial pooling effect sizes that can modify these global effects. While Figure 9 does show regional differences are prevalent, the global parameters explain most of the variation in the reduction of *R_t_*.

### 2.2 Attack rates

Despite Italy having the largest number of deaths attributable to COVID-19 in Europe, the estimated attack rate (percentage of the population that has been infected) is still relatively low across all regions (Table 1). We estimate the highest attack rates in Lombardy and in Aosta Valley (13.30% [10.72%-16.69%] and 11.07% [8.38%-14.73%], respectively) with many regions having an average attack rates of less than 1%. Even in the highest mortality regions, the attack rates are far from the herd immunity threshold (~ 70% assuming an *R*_0_ = 3). Simulating 8 weeks into the future, using a scenario of a 20% return to pre-lockdown levels of mobility, we estimate the highest attack rates to be in Piedmont with 19.64% [13.22%-28.05%], Lombardy with 13.79% [11.12%-17.31%] and Veneto with 12.90% [7.65%-20.18%]. In the scenario of a larger 40% return to pre-lockdown levels of mobility, over 8 weeks the attack rate in Piedmont is estimated to become 54.18% [41.71%-65.52%], followed by Tuscany with 41.71% [21.06%-62.24%] (see Table 3).

### 2.3 Transmission intensity estimates over time

Figure 6 shows the basic reproduction number (*R*_0_) and effective reproduction number (*R****_t_***) as of 1*st* of May 2020. The posterior mean basic reproduction number is 3.4[2.6 – 4.3] and in line with that previously reported [8].The posterior mean of the current reproduction number is below one, and is significantly so for the majority of regions. These results provide strong evidence that the major NPIs implemented have universally controlled the epidemic across all of Italy.

**Figure 4:**
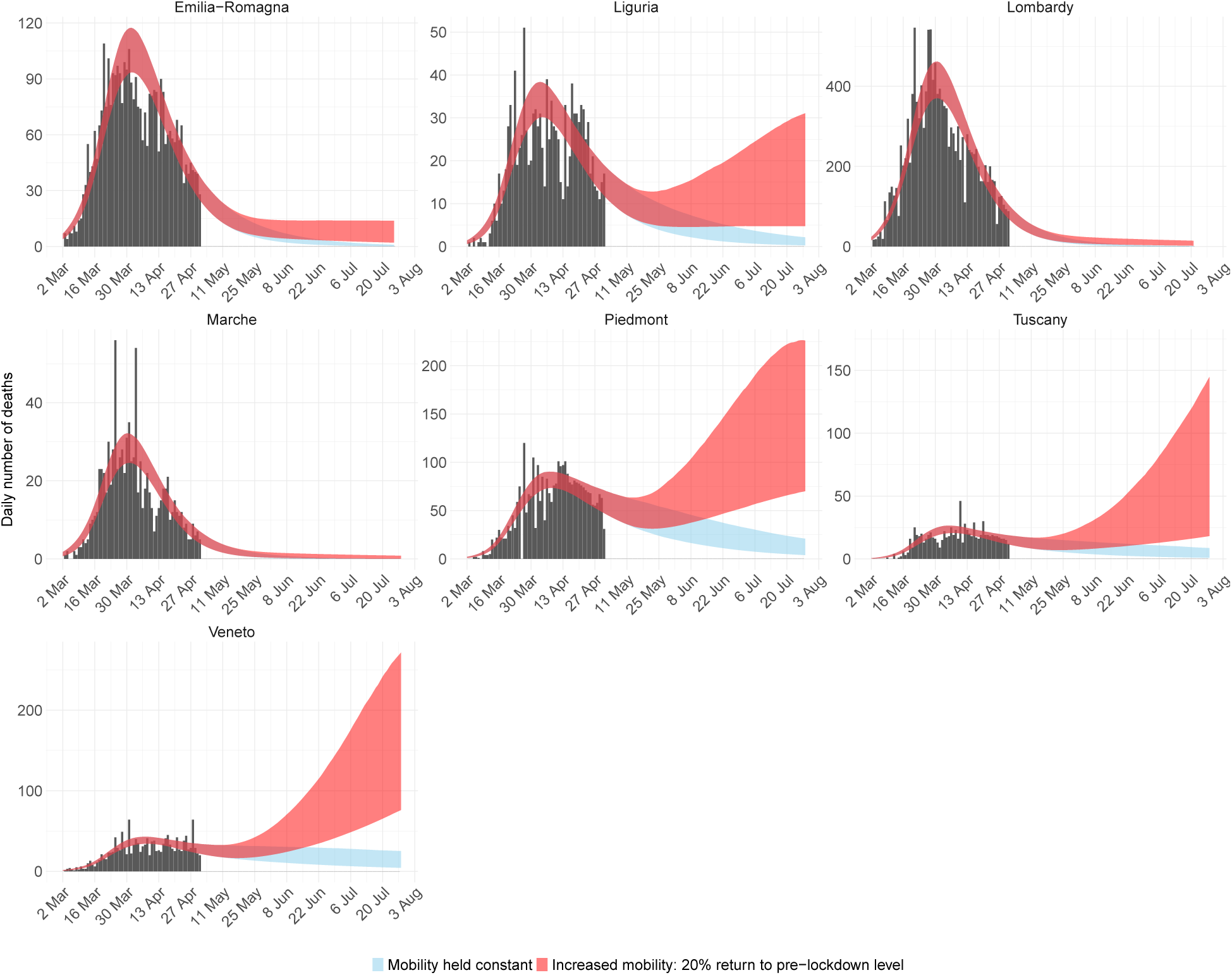
Deaths for the 7 regions with highest mortality in Italy. Black bars are the data, blue ribbons is the 95% credible interval forecast scenario were mobility stays at lockdown levels, and red is the 95% credible interval forecast scenario where mobility returns by 20% to pre-lockdown levels.

**Figure 5:**
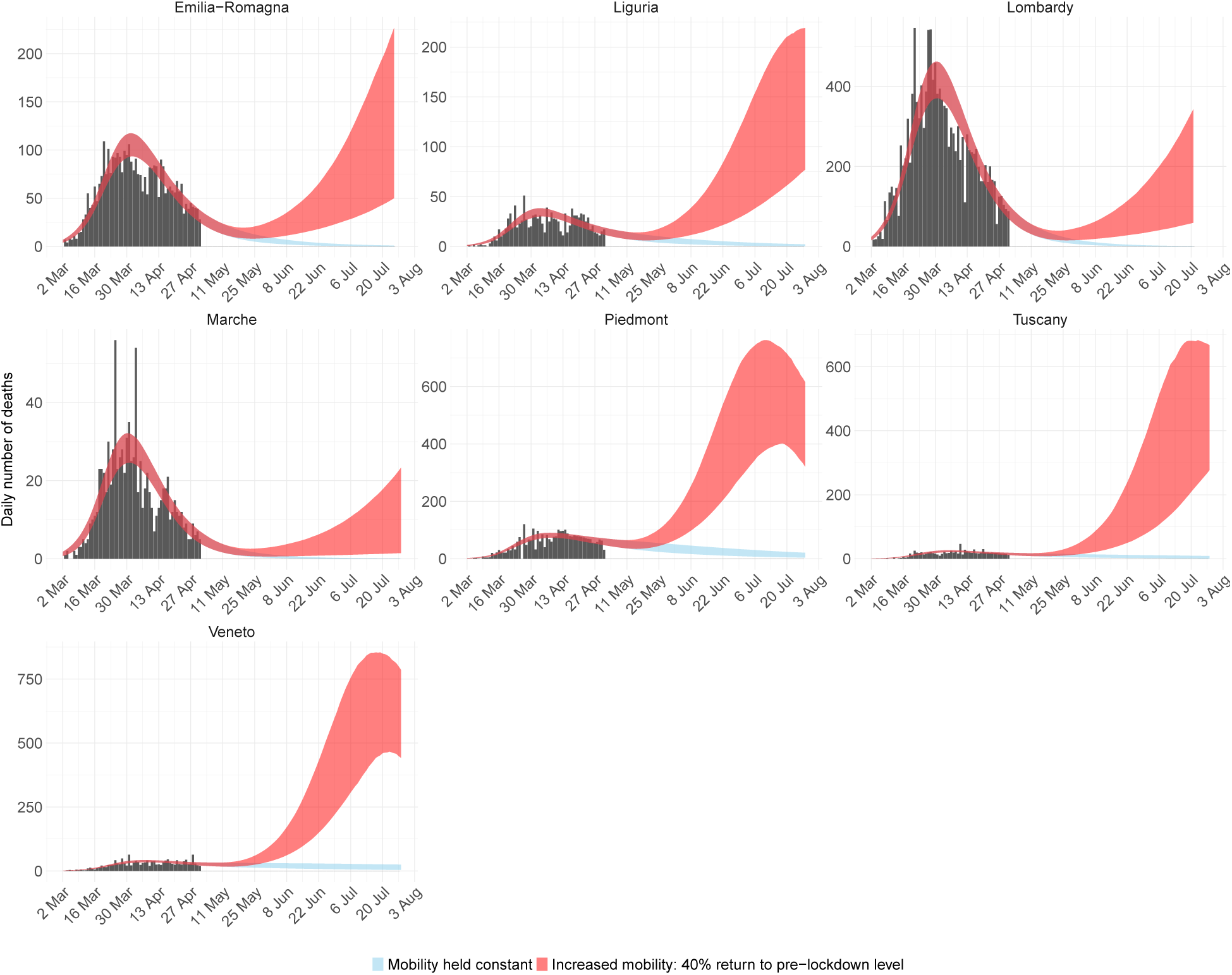
Deaths for the 7 regions with highest mortality in Italy. Black bars are the data, blue ribbon is the 95% credible interval forecast scenario were mobility stays at lockdown levels, and red is the 95% credible interval forecast scenario where mobility returns by 40% to pre-lockdown levels.

**Figure 6:**
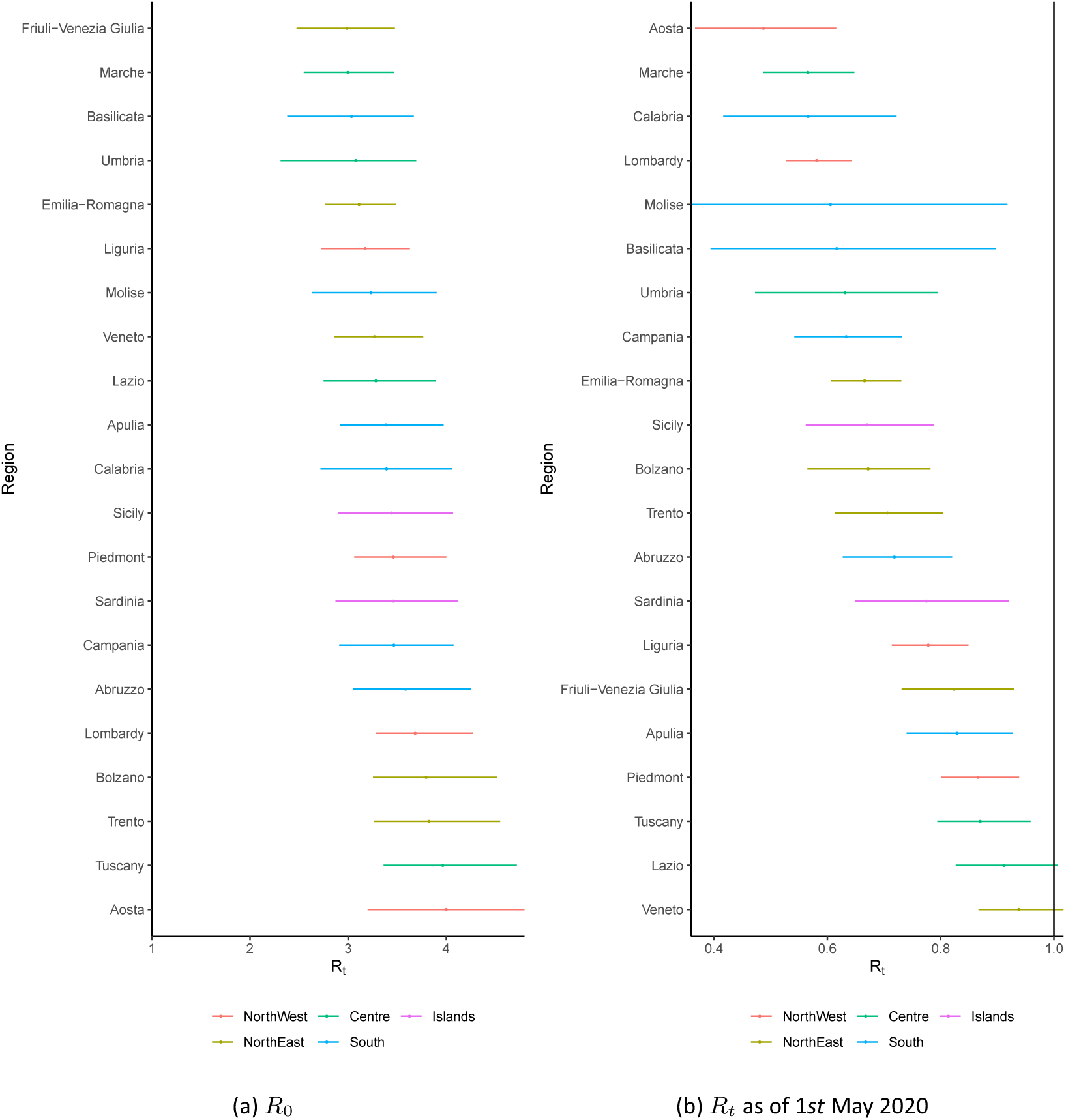
Regional estimates of *R_t_*. Figure (a) shows the basic reproduction number *R_0_* and (b) shows the estimate of *R_t_* as of 1*st* May 2020. The colours indicate the 5 Italian macro regions.

Figures 7 and 8 show the model fits for the 7 regions with the highest mortality of COVID-19 in Italy. Visually, there is a strong correspondence between large drops in *R_t_* and intervention timings. This suggests that interventions have had a strong effect on mobility, which our model then translates into effects on transmission intensity. From the mobility data, there are clear day-of-the-week fluctuations that affect transmission, but these fluctuations are small compared to the overall reductions in mobility. For all of the 7 regions with the highest mortality rates we see a large reduction in infections, with the turning point coincident with the onset of interventions and the subsequent reductions on mobility. While we estimate the daily numbers of infections to be in rapid decline, due to the lag between infections and deaths, more deaths will occur in the next weeks.

### 2.4 Future simulation scenarios

The primary mechanism driving dynamics in our model is *R****_t_***, which is parameterised by mobility. Using our model, jointly fitted to all regions in Italy, we are able to simulate forwards 8 weeks with hypothetical scenarios where mobility increases. We do not differentiate what causes these increases in mobility but it stands to reason they would occur from a relaxation of NPIs and changes in behaviour. We also note that other mechanisms aside from mobility can increase *R_t_* and would yield in the same result.

We choose three scenarios (a) **constant mobility** in which mobility remains at current lockdown levels for 8 weeks, (b) 20% **return to pre-lockdown mobility** and (c) 40% **return to pre-lockdown mobility**.

Scenarios (b) and (c) are calculated using a weighted average between the current mobility and the nominal pre-lockdown level. Thus, for example, in scenario (b), 20% of the weight is on the nominal pre-lockdown level and 80% on the current mobility. Scenario (a) is equivalent to a 0% return to prelockdown mobility.

Figures 7 and 8 show the estimated increases in *R_t_* due to a 40% return to pre-lockdown mobility. A 40% return represents a reasonably large change in mobility and for many regions shifts *R_t_* just above 1. The result of this increase in *R_t_* manifests in a rise in the number of daily infections and deaths. Figures 4 and 5 show the scenarios of 20% and 40% returns to pre-lockdown mobility. In the constant mobility scenario we predict a continued reduction in deaths, however in the 20% and 40% scenarios, while initially deaths may continue to decrease, there will eventually be a resurgent epidemic that, without accounting for additional interventions, may be larger in size than the first wave.

**Figure 7:**
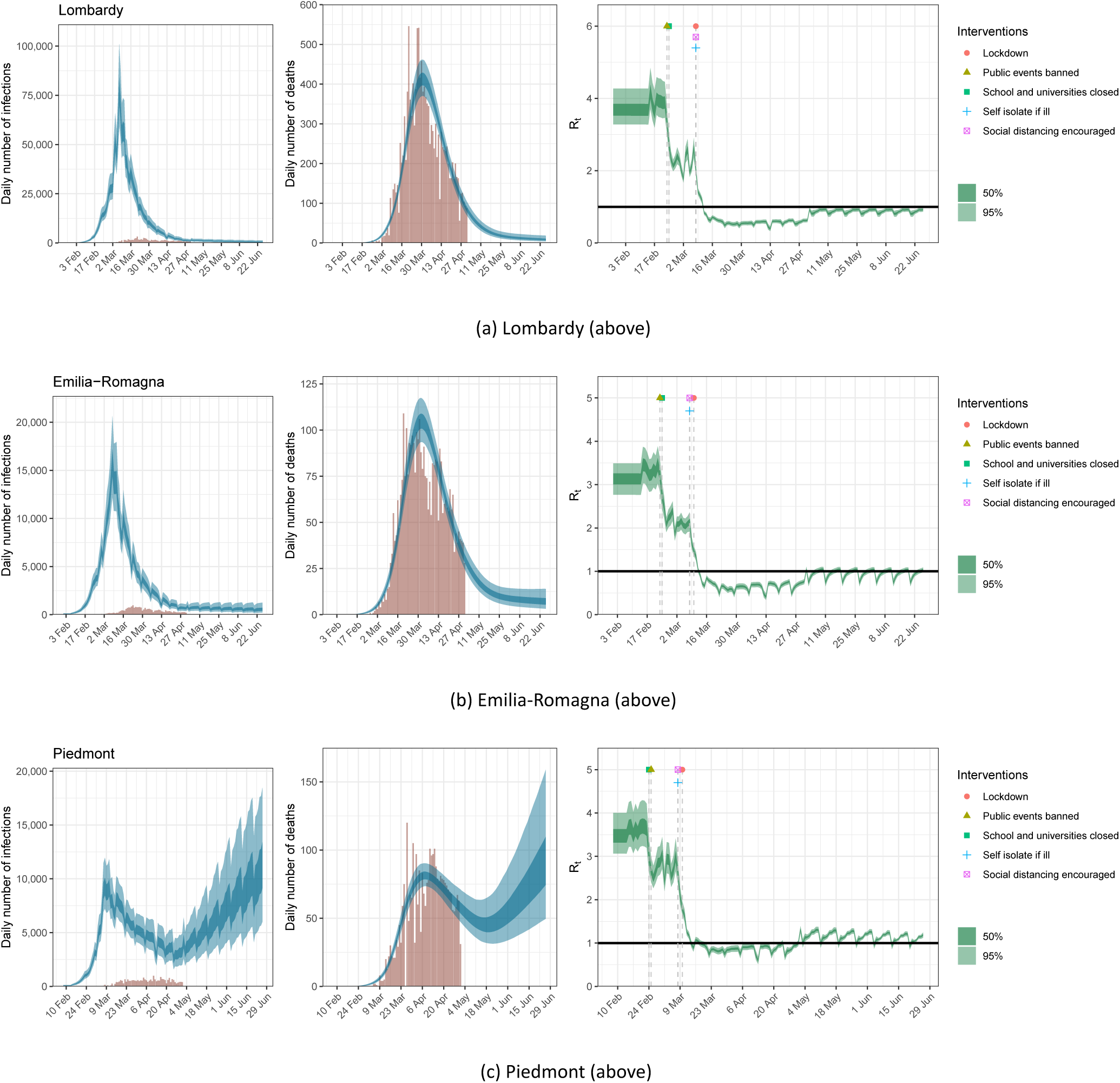
Estimates of infections, deaths and R_t_ for Lombardy, Emilia-Romagna and Piedmont under the scenario of a 20% return to pre-lockdown levels of mobility. Left: daily number of infections, brown bars are reported cases, blue bands are predicted infections, dark blue 50% credible interval (CI), light blue 95% CI. The number of daily infections estimated by our model immediately responds to changes in mobility, as we assume that all infected people become immediately less or more infectious. If the R_t_ is above 1, the number of infections will starts growing again. Middle: daily number of deaths, brown bars are reported deaths, blue bands are predicted deaths, CI as in left plot. Right: time-varying reproduction number R_t_, dark green 50% CI, light green 95% CI. Icons are interventions shown at the time they occurred.

**Figure 8:**
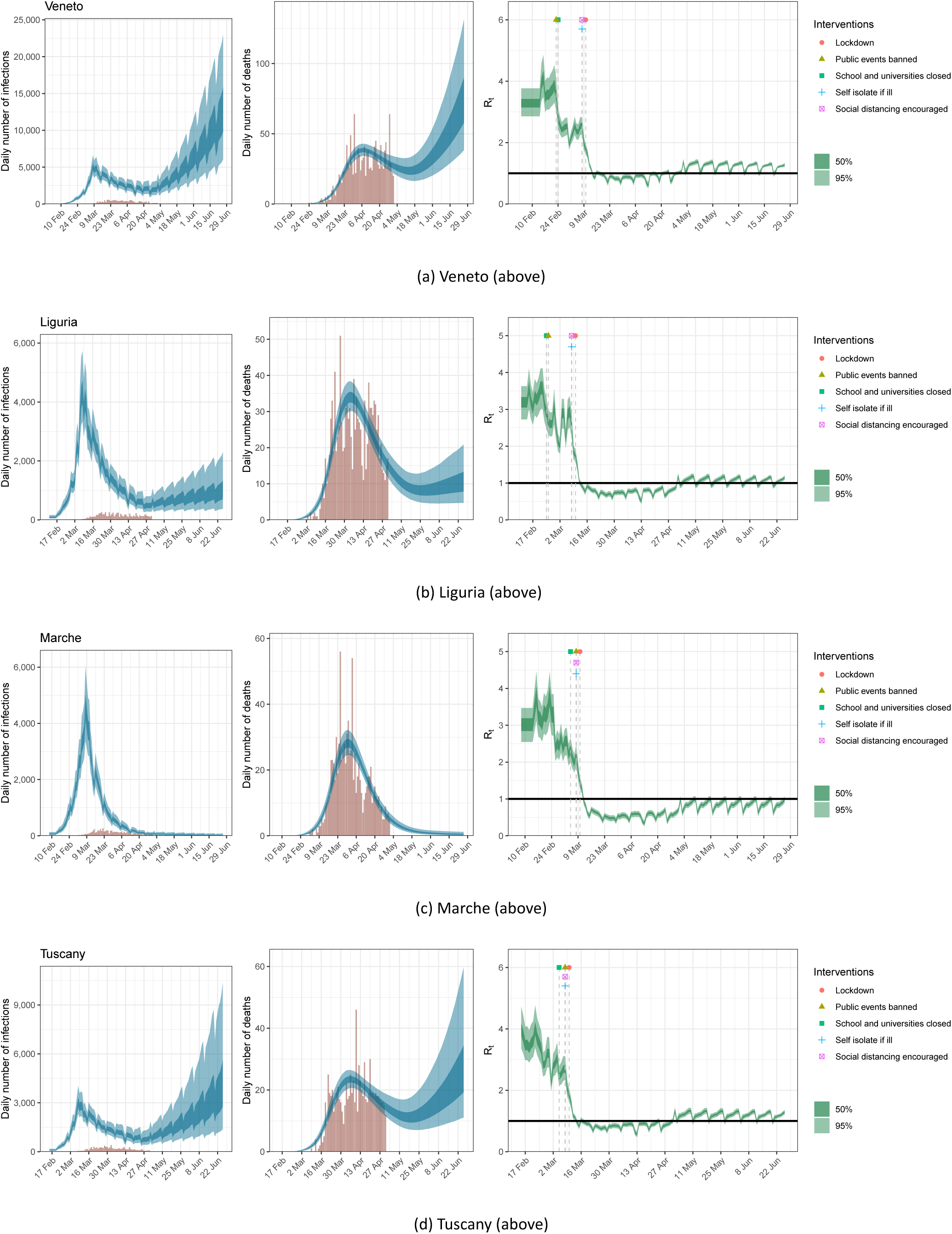
Estimates of infections, deaths and R_t_ for Veneto, Liguria, Marche and Tuscany under the scenario of a 20% return to pre-lockdown levels of mobility; same plots as in Figure 7.

**Figure 9:**
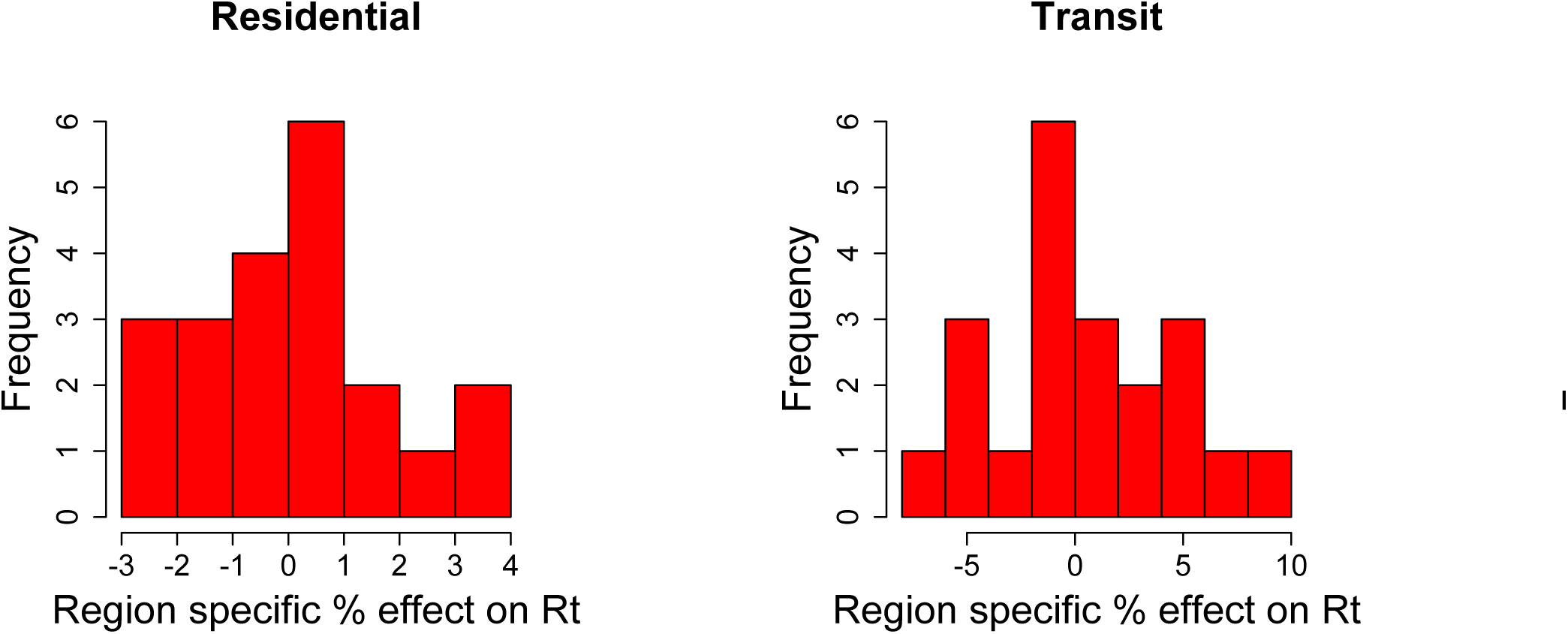
Partial pooling effect sizes for all three mobility dimensions

### 2.5 Deaths averted under future simulation scenarios

Using our simulated scenarios we can calculate the deaths averted by keeping mobility at current levels. Table 2 shows the deaths averted under the 20% and 40% return to pre-lockdown mobility scenarios and no other intervention is put in place. Under the 20% scenario we estimate the total number of excess deaths to be between 3,000 and 5,000, and under the 40% scenario the total number of excess deaths would be between 10,000 and 23,000 (see Table 2). The deaths averted are largest in regions currently experiencing major epidemics; the reason they rebound to such a large extent is driven by a large number of ongoing infections. If more time is spent under current lockdown mobility levels before increases occur, the number of deaths averted is likely to be considerably lower in both scenarios. It should be noted that in our model we do not account for cross-region movement, which, given increased mobility, is likely to increase infections and subsequently deaths, in regions not experiencing major epidemics.

**Table 2:** Number of deaths averted assuming mobility at the lockdown levels for 8 weeks from 1st May 2020 (scenario a) compared to a 20% return to pre-lockdown levels (scenario b) or a 40% return to pre-lockdown levels (scenario c) with mean and the [95% credible interval].

| Region | deaths averted if lockdown maintained<br>vs. a 20% return to pre-lockdown mobility | deaths averted if lockdown maintained<br>vs. a 40% return to pre-lockdown mobility |
| --- | --- | --- |
| Abruzzo | 35 [10-88] | 200 [60-490] |
| Basilicata | 2 [0-13] | 11 [0-75] |
| Calabria | 2 [0-7] | 10 [1-45] |
| Campania | 14 [3-35] | 82 [20-220] |
| Emilia-Romagna | 130 [60-230] | 650 [300-1,100] |
| Friuli-Venezia Giulia | 56 [20-130] | 260 [80-610] |
| Lazio | 330 [100-640] | 1700 [700-3,300] |
| Liguria | 160 [80-290] | 760 [400-1,400] |
| Lombardy | 190 [90-350] | 1,100 [500-2,000] |
| Marche | 9 [3-22] | 49 [20-110] |
| Molise | 2 [0-14] | 10 [0-72] |
| Bolzano | 10 [2-28] | 57 [10-160] |
| Trento | 23 [6-56] | 120 [30-290] |
| Piedmont | 1,300 [700-2,100] | 5,600 [3,000-8,700] |
| Apulia | 140 [50-310] | 790 [300-1,700] |
| Sardinia | 30 [5-96] | 170 [30-540] |
| Sicily | 14 [3-41] | 85 [20-260] |
| Tuscany | 370 [200-720] | 1,800 [800-3,600] |
| Umbria | 3 [0-12] | 15 [1-66] |
| Aosta Valley | 1 [0-2] | 3 [0-14] |
| Veneto | 930 [500-1,600] | 4,100 [2,000-6,600] |
| Total | 3,700 [3,000-5,000] | 18,000 [10,000-23,000] |

**Table 3:** Attack Rate after 8 weeks from 1st May 2020 if mobility returns to a 20% pre-lockdown levels (scenario b) or a 40% return to pre-lockdown levels (scenario c) with mean and the [95% credible interval]

| Regions | 20% return to pre-lockdown mobility | 40% return to pre-lockdown mobility |
| --- | --- | --- |
| Abruzzo | 3.83% [2.49%-6.53%] | 18.57% [5.91%-42.30%] |
| Basilicata | 0.84% [0.27%-3.35%] | 6.06% [0.34%-47.89%] |
| Calabria | 0.53% [0.38%-0.79%] | 1.27% [0.44%-5.07%] |
| Campania | 0.85% [0.64%-1.17%] | 2.61% [0.91%-7.49%] |
| Emilia-Romagna | 8.71% [6.99%-11.00%] | 13.14% [9.53%-18.33%] |
| Friuli-Venezia Giulia | 4.64% [2.68%-8.82%] | 19.26% [6.48%-40.57%] |
| Lazio | 4.93% [2.15%-9.74%] | 31.57% [12.52%-54.01%] |
| Liguria | 10.72% [7.63%-15.27%] | 31.08% [18.11%-46.28%] |
| Lombardy | 13.79% [11.12%-17.31%] | 16.66% [13.06%-21.37%] |
| Marche | 5.50% [4.36%-6.88%] | 6.62% [4.92%-9.13%] |
| Molise | 1.21% [0.34%-6.52%] | 7.69% [0.43%-57.23%] |
| Bolzano | 6.84% [5.08%-9.69%] | 17.13% [7.06%-39.45%] |
| Trento | 10.23% [7.58%-14.63%] | 26.80% [12.50%-49.54%] |
| Piedmont | 19.64% [13.22%-28.05%] | 54.18% [41.47%-65.52%] |
| Apulia | 3.47% [1.62%-7.38%] | 25.14% [8.51%-48.77%] |
| Sardinia | 2.12% [0.80%-6.59%] | 17.69% [2.39%-51.95%] |
| Sicily | 0.69% [0.48%-1.16%] | 3.67% [0.78%-13.01%] |
| Tuscany | 8.26% [4.14%-15.52%] | 41.71% [21.06%-62.24%] |
| Umbria | 0.86% [0.54%-1.56%] | 2.69% [0.65%-12.14%] |
| Aosta | 11.22% [8.49%-14.83%] | 12.53% [8.97%-18.64%] |
| Veneto | 12.90% [7.65%-20.18%] | 43.90% [28.69%-58.70%] |

## 3 Conclusions

In this report we use a semi-mechanistic Bayesian hierarchical model fitted to sub-national death data for Italy. We parameterise the reproduction number, a fundamental measure of transmission intensity, as a function of an individual’s mobility. We show that mobility, both visually and statistically, is associated with the onset and timing of major NPIs. Using our model, we estimate that the average reproduction numbers in all regions across Italy is currently below 1, suggesting that the major interventions implemented by the Italian government have controlled transmission and averted a major health catastrophe. We show that despite the large number of deaths attributable to COVID-19, the attack rates are far lower than required for herd immunity. Simulating 8 weeks into the future, we estimate that, if mobility remains the same, there will be a continued reduction in deaths and the epidemic will be suppressed. However, returns to pre-lockdown mobility of 20% or 40% from current levels may lead to a resurgence of the epidemic with more deaths than the current wave in the absence of additional interventions.

Our modelling framework is novel in that we infer a latent function for infections, and, to the best of our knowledge for the first time, parameterise *R_t_* using mobility data. The use of mobility data as a proxy for the time spent in day-to-day activities does not capture individual-level measures such as case isolation and only partially captures the impact of other interventions such as school and university closure. When simulating future scenarios we have not accounted for the impact of social distancing measures in public transport and public space, alongside the mandatory use of personal protective equipment (PPE). The cost benefit trade off between the implementation of new interventions and the relaxation of current NPIs is unknown, and will depend on the effectiveness of this new set of interventions, behaviour, adherence to the recommendations and the correct use of the personal protective equipment. Given that interventions, such as extensive testing, contact tracing and social distancing are going to be implemented, our estimates can be viewed as being pessimistic. On the other hand, simulating 20% and 40% increase in mobility over the next 8 weeks is likely a conservative scenario. Our model uses the official deaths counts to estimate changes in transmission intensity. We did not use the reported number of confirmed COVID-19 cases because of potential biases arising from changes in the case definition and testing strategy adopted during the epidemic across the regions, which would be hard to correct for. As more information on new interventions are introduced we will include them in our modelling framework.

Our results suggest that transmission, as well as mobility, need to be closely monitored in the future weeks and months. To date, it is hard to predict the extent to which new interventions will be able to maintain *R_t_* around 1 once the currently implemented NPIs are relaxed. The success of the new interventions such as social distancing on public transports and the use of personal protective equipment depends on population behaviour, adherence to recommendations, the effectiveness and correct use of the equipment as well as on the enhanced and timely monitoring of SARS-CoV-2 transmission. Because increases in the number of hospitalisations and deaths lag behind increases in transmission intensity, the control of a future potential resurgence in transmission relies on the early identification and isolation of infections and on the timely suppression of local clusters of infection. Enhanced disease surveillance via swab testing and contact tracing allows to identify infections early and to timely monitor changes in transmission intensity and is key to compensate for the risk of resurgence in transmission that may occur following the increase in mobility that is likely to be observed once that the current NPIs will be relaxed.

## 4 Data

Our model utilizes daily real-time death data provided by the Italian Civil Protection (publicly available at https://github.com/pcm-dpc/COVID-19) for the 20 Italian regions. For the Trentino Alto-Adige region, we report the results for the provinces of Trento and Bolzano separately, following the format of the death data provided by the Italian Civil Protection. For population counts, we use publicly available age-stratified counts from ISTAT (“Popolazione residente comunale per sesso anno di nascita e stato civile”, from https://www.istat.it).

Mobility data have been obtained from the Google Mobility Report (google.com/covid19/mobility/) which provides data on movement in Italy by region and highlights the percent change in visits to:

- Grocery & pharmacy: Mobility trends for places like grocery markets, food warehouses, farmers markets, specialty food shops, drugstores, and pharmacies.
- Parks: Mobility trends for places like local parks, national parks, public beaches, marinas, dog parks, plazas, and public gardens.
- Transit stations: Mobility trends for places like public transport hubs such as subway, bus, and train stations.
- Retail & recreation: Mobility trends for places like restaurants, cafes, shopping centers, theme parks, museums, libraries, and movie theaters.
- Residential: Mobility trends for places of residence.
- Workplaces: Mobility trends for places of work.

The mobility data show the length of stay at different places compared to a baseline. It is therefore relative, i.e mobility of −50% means that, when compared to pre COVID-19, individuals are engaging in a given activity 50% less.

We also catalogue data on the nature and type of major NPIs. We referred to government as well as official public health division webpages to identify the recommendations and laws being issued by the central government and local public health authorities. We collected the following:

- School closure ordered: This intervention refers to nationwide extraordinary school closures which in most cases refer to both primary and secondary schools closing (for most regions this also includes the closure of other forms of higher education or the advice to teach remotely). The date of the school closure is taken to be the effective date when the schools started to be closed (if this was on a Monday, the date used was the one of the previous Saturdays as pupils and students effectively stayed at home from that date onwards).
- Case-based measures: This intervention comprises strong recommendations or laws to the general public and primary care about self-isolation when showing COVID-19-like symptoms. These also include nationwide testing programs where individuals can be tested and subsequently self-isolated. Our definition is restricted to official advice to all individuals or to all primary care. These do not include containment phase interventions such as isolation if travelling back from an epidemic region such as China.
- Public events banned: This refers to banning all public events of more than 100 participants such as sports events.
- Social distancing encouraged: As one of the first interventions against the spread of the COVID-19 pandemic, the central government and many regions published advice on social distancing including the recommendation to work from home wherever possible and reduce the use of public transport and all other non-essential contacts. The dates used are those when social distancing has officially been recommended; the advice may include maintaining a recommended physical distance from others.
- Lockdown decreed: There are several different scenarios that the media refers to as lockdown. As an overall definition, we consider regulations/legislations regarding strict face-to-face social interaction: including the banning of any non-essential public gatherings, closure of educational and public/cultural institutions, ordering people to stay home apart from essential tasks. We include special cases where these are not explicitly mentioned on government websites but are enforced by the police. The dates used are the effective dates when these legislations have been implemented. We note that lockdown encompasses other interventions previously implemented.

The mobility data together with the intervention timings are shown in Figure 2.

## 5 Methods

In a previous report [4], we introduced a new Bayesian framework for estimating the transmission intensity and attack rate (percentage of the population that has been infected) of COVID-19 from the reported number of deaths. This framework uses the time varying reproduction number *R_t_* to inform a latent function for infections, and then these infections, together with probabilistic lags, are calibrated against observed deaths. Observed deaths, while still susceptible to under reporting and delays, are more reliable than the reported number of confirmed cases, although the early focus of most surveillance systems on cases with reported travel histories to China may have missed some early deaths. Changes in testing strategies during the epidemic mean that the severity of confirmed cases as well as the reporting probabilities changed in time and may thus have introduced bias in the data.

In this report, we adapt our original Bayesian semi-mechanistic model of the infection cycle to the 20 Italian regions. We infer plausible upper and lower bounds (Bayesian credible intervals) of the total populations infected (attack rates) and the reproduction number over time (*R****_t_***). In our framework we parameterise *R_t_* as a function of Google mobility data. We fit the model jointly to COVID-19 data from all regions to assess whether there is evidence that changes in mobility have so far been successful at reducing *R_t_* below 1. Our model is a partial pooling model, where the effect of mobility is shared but region-specific modifiers can capture differences and idiosyncrasies among the regions. We then simulate forwards using a simple assumption that mobility returns to 20% or 40% pre-lockdown levels of mobility from the latest lockdown levels and explore the impact of increased mobility on transmission intensity, infections and deaths.

We note that future directions should focus on embedding mobility in realistic contact mechanisms to establish a closer relationship to transmission.

### Model specifics

We observe daily deaths *D_t_,_m_* for days *t* ∈ {1, …, *n*} and regions *m* ∈ {1, …,*M*}. These daily deaths are modelled using a positive real-valued function 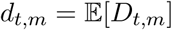 that represents the expected number of deaths attributed to COVID-19. The daily deaths *D_t_,_m_* are assumed to follow a negative binomial distribution with mean *d_t_,_m_* and variance 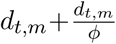, where *ψ* follows a positive half normal distribution, i.e.

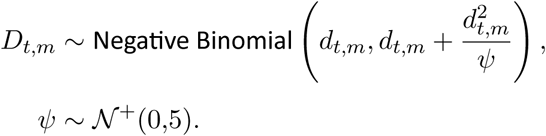

Here, 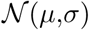 denotes a normal distribution with mean *μ* and standard deviation *σ*. We say that *X* follows a positive half normal distribution 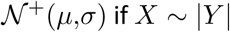, where 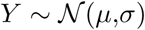.

To mechanistically link our function for deaths to our latent function for infected cases, we use a previously estimated COVID-19 infection fatality ratio (ifr, probability of death given infection) together with a distribution of times from infection to death *π*. Details of this calculation can be found in [16, 17]. From the above, every region has a specific mean infection fatality ratio ifr*_m_* (see Table 1). To incorporate the uncertainty inherent in this estimate we allow the ifr*_m_* for every region to have additional noise around the mean. Specifically we assume

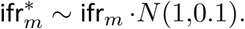

Using estimated epidemiological information from previous studies[16, 17], we assume the distribution of times from infection to death *π* (infection-to-death) to be

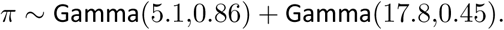

The expected number of deaths *d_t_,_m_*, on a given day *t*, for region, *m*, is given by the following discrete sum:

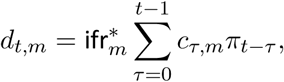

where *c_τ_,_m_* is the number of new infections on day *τ* in region *m* and where *π* is discretized via 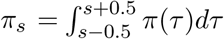 for *s* = 2,3,…, and 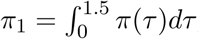, where 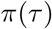 is the density of *π*.

The true number of infected individuals, *c*, is modelled using a discrete renewal process. We specify a generation distribution *g* with density *g*(*τ*) as:

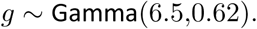

Given the generation distribution, the number of infections *c_t_,_m_* on a given day *t*, and region, *m*, is given by the following discrete convolution function:

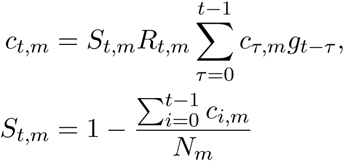

where, similar to the probability of death function, the generation distribution is discretized by 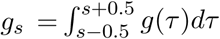 for *s =* 2,3,…, and 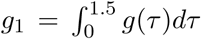. The population of region *m* is denoted by *N_m_*. We include the adjustment factor 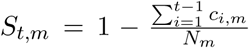 to account for the number of susceptible individuals left in the population.

We parametrise *R_t_,_m_* as a linear function of the relative change in time spent (from a baseline) across three (*k* = 3) Google mobility dimensions: residential, transit station and an average of retail and recreation, groceries and pharmacies, parks, and workplaces. The reason for taking an average was that these dimensions were extremely collinear. The effect of mobility on transmission is assumed to be multiplicative. *R_t_,_m_* is therefore a function of the mobility indicator *I_k,t,m_* in place at time *t* in region *m*:

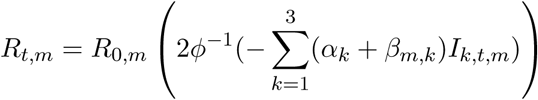

where *ϕ*^-1^ is the inverse logit or sigmoid function. The impacts *α_k_* are shared between all *M* regions and *β_m_,_k_* allows for region specific effects. This model is therefore a partial pooling model. The prior distribution for the shared coefficients were chosen to be

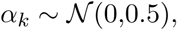

and the prior distribution for the pooled coefficients were chosen to be

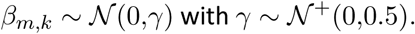

The prior distribution for *R*_0_,*_m_*[8] was chosen to be

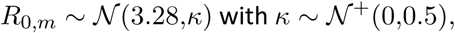

where *k* is the same among all regions.

We assume that seeding of new infections begins 30 days before the day after a region has cumulatively observed 10 deaths. From this date, we seed our model with 6 sequential days of an equal number of infections: 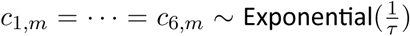, where *τ* ~ Exponential(0.03). These seed infections are inferred in our Bayesian posterior distribution.

We estimated parameters jointly for all regions in a single hierarchical model. Fitting was done in the probabilistic programming language Stan[2] using an adaptive Hamiltonian Monte Carlo (HMC) sampler.

## Data Availability

Our model utilizes daily real-time death data provided by the Italian Civil Protection (publicly available at https://github.com/pcm-dpc/COVID-19) for the 20 Italian regions. For the Trentino Alto-Adige region, we report the results for the provinces of Trento and Bolzano separately, following the format of
the death data provided by the Italian Civil Protection. For population counts, we use publicly available age-stratified counts from ISTAT (”Popolazione residente comunale per sesso anno di nascita e stato civile”, from https://www.istat.it). Mobility data have been obtained from the Google Mobility Report (google.com/covid19/mobility/) which provides data on movement in Italy by region

https://www.istat.it

## Acknowledgements

We would like to thank Amazon AWS and Microsoft Azure for computational credits. We would like to thank the Stan Development team for their constant support.

### 6 Appendix

#### 6.1 Results for the Italian regions not shown in the main text

**Figure 10:**
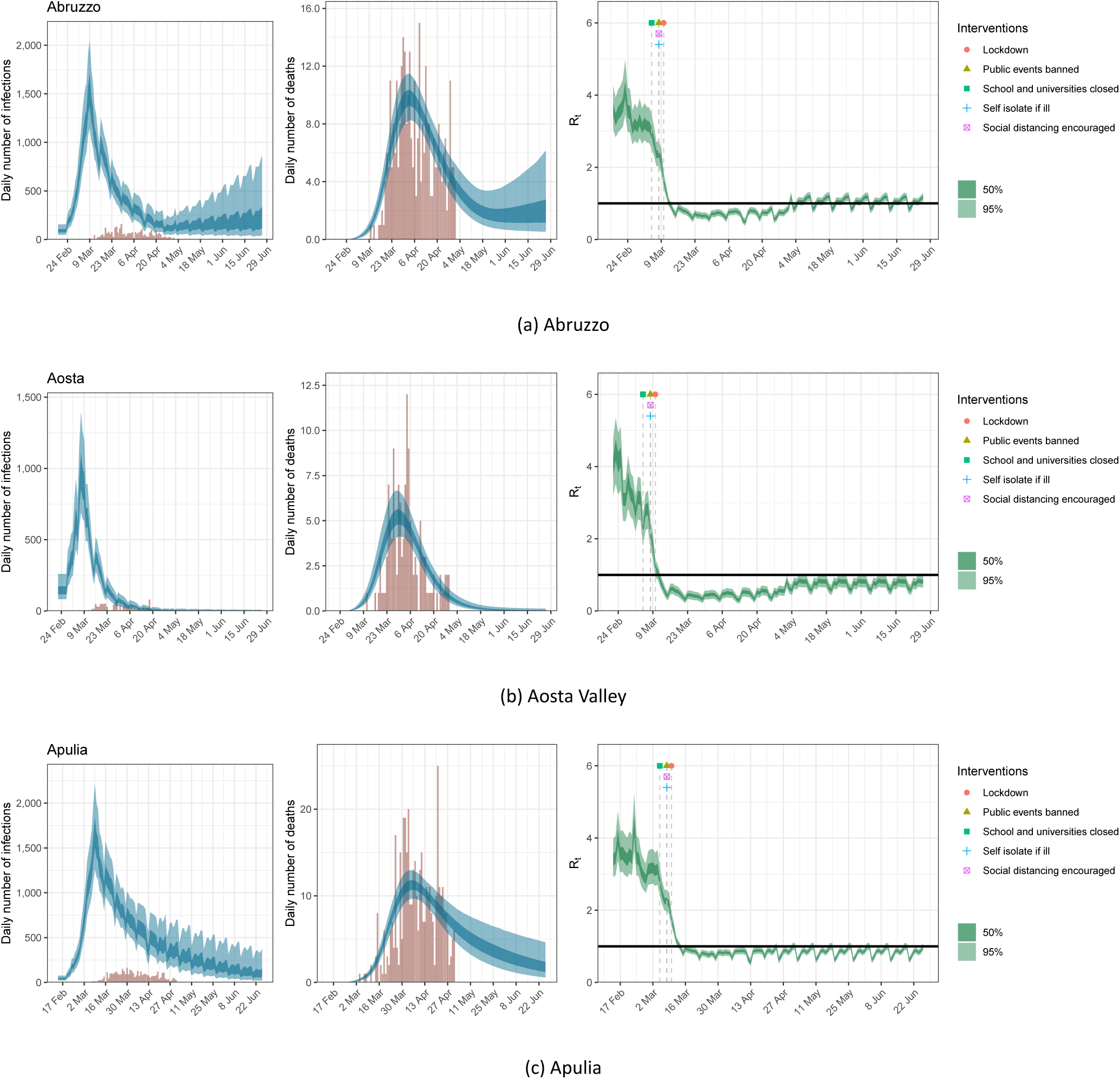
Estimates of infections, deaths and R_t_ for Abruzzo, Aosta Valley and Apulia under the scenario of a 20% return to pre-lockdown levels of mobility; same plots as in Figure 7.

**Figure 11:**
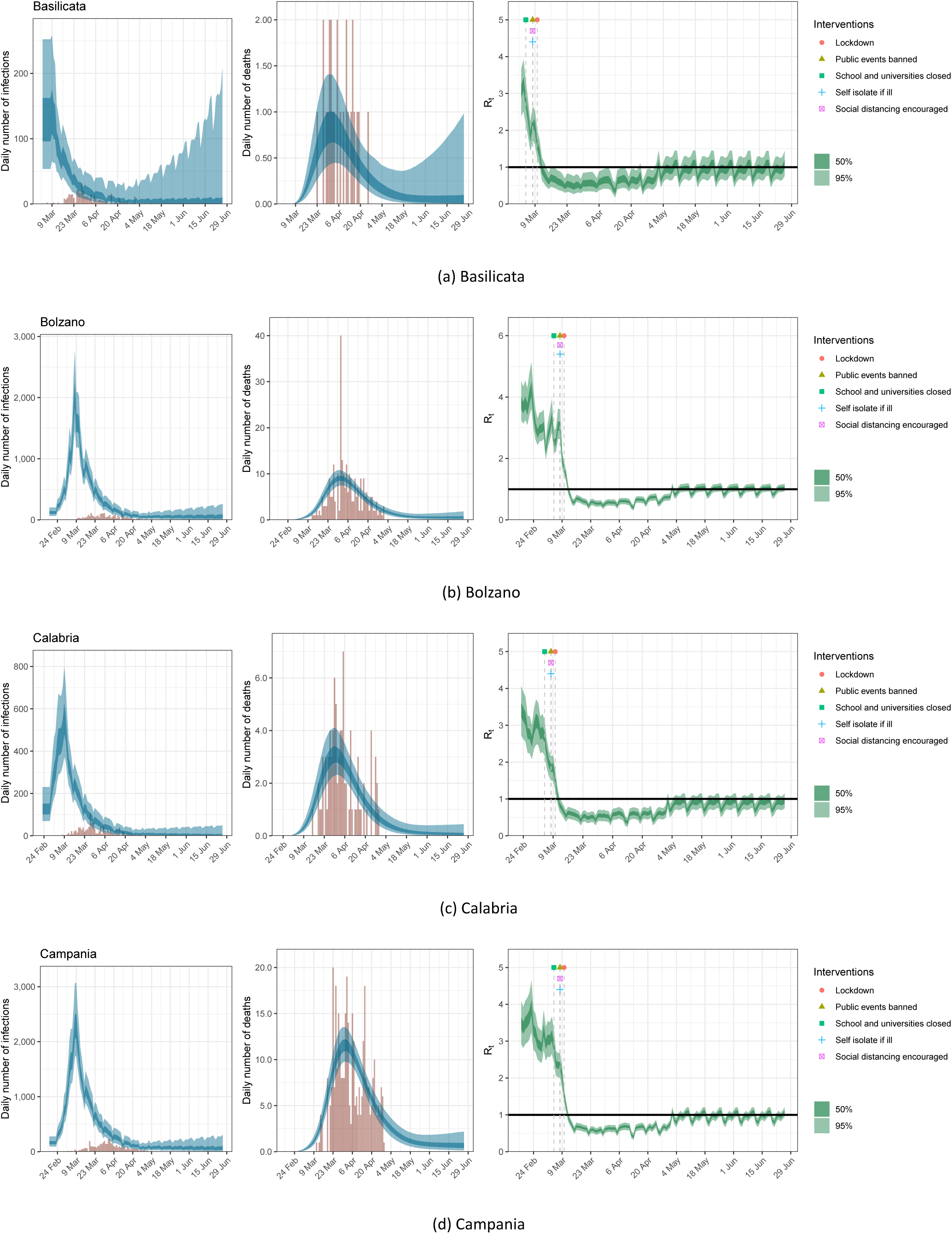
Estimates of infections, deaths and R_t_ for Basilicata, Bolzano, Calabria and Campania under the scenario of a 20% return to pre-lockdown levels of mobility; same plots as in Figure 7.

**Figure 12:**
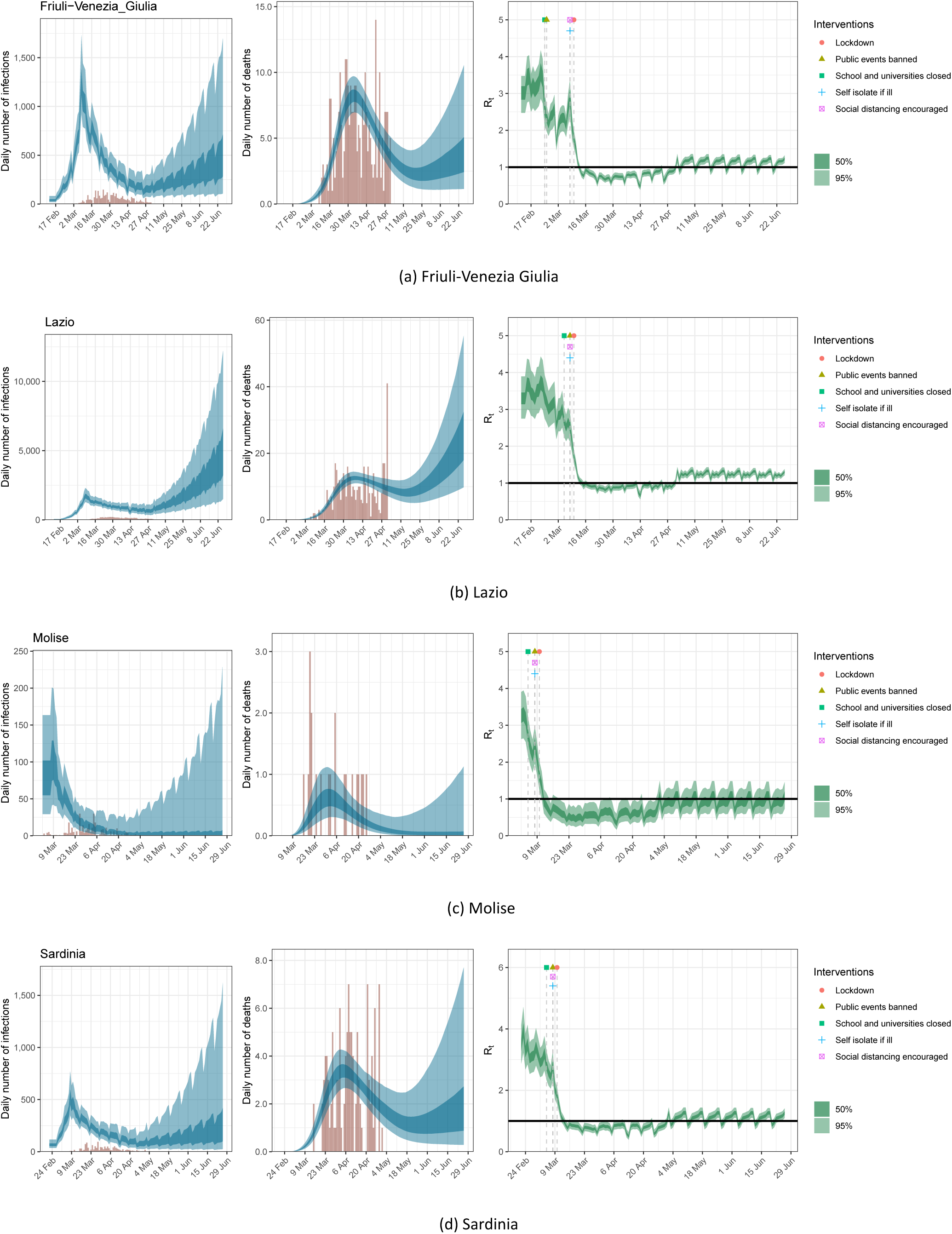
Estimates of infections, deaths and R_t_ for Friuli-Venezia Giulia, Lazio, Molise and Sardinia under the scenario of a 20% return to pre-lockdown levels of mobility; same plots as in Figure 7.

**Figure 13:**
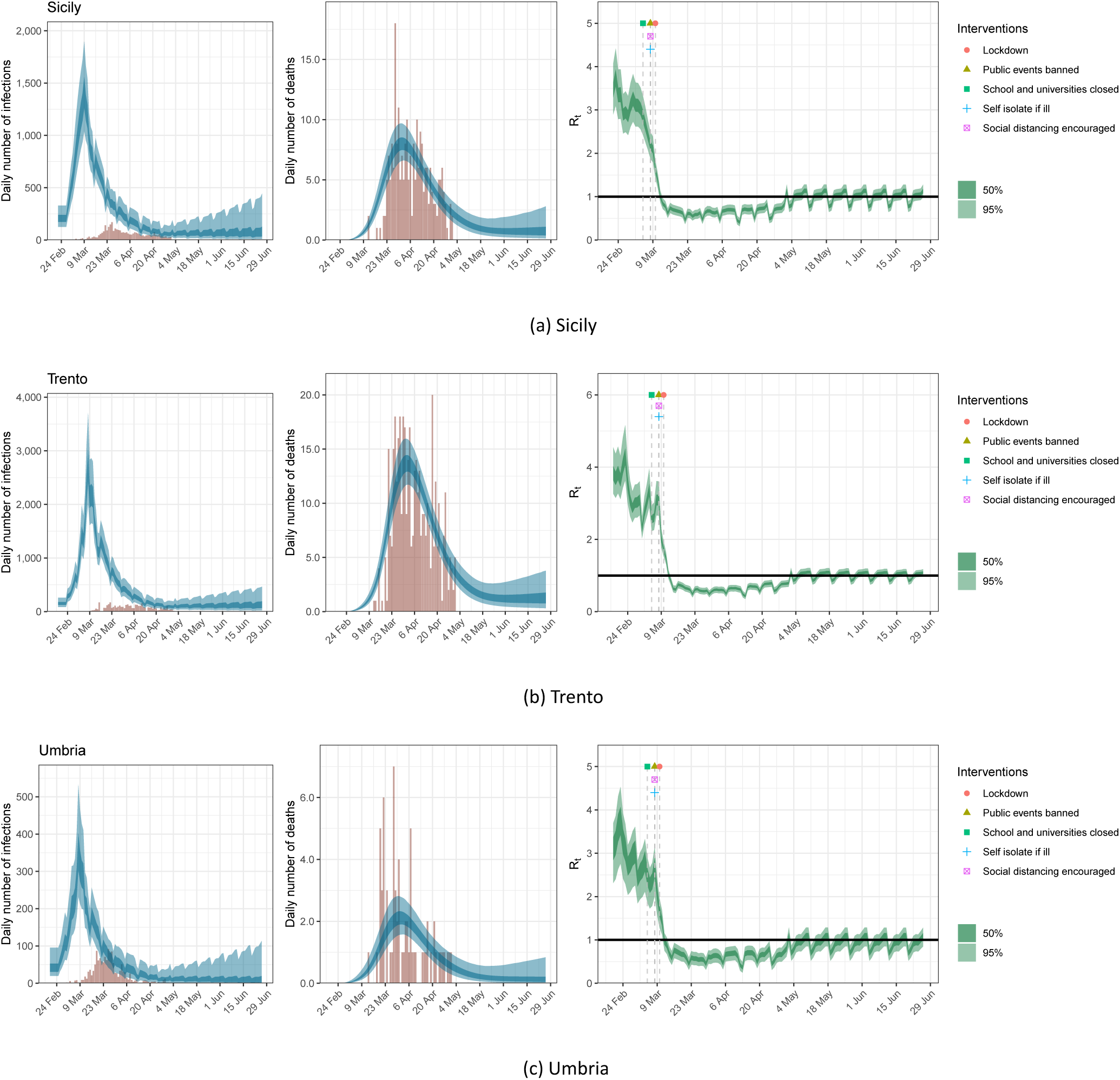
Estimates of infections, deaths and R_t_ for Sicily, Umbria and Trento under the scenario of a 20% return to pre-lockdown levels of mobility; same plots as in Figure 7.

#### 6.2 Scenarios for the Italian regions not included in the main text

**Figure 14:**
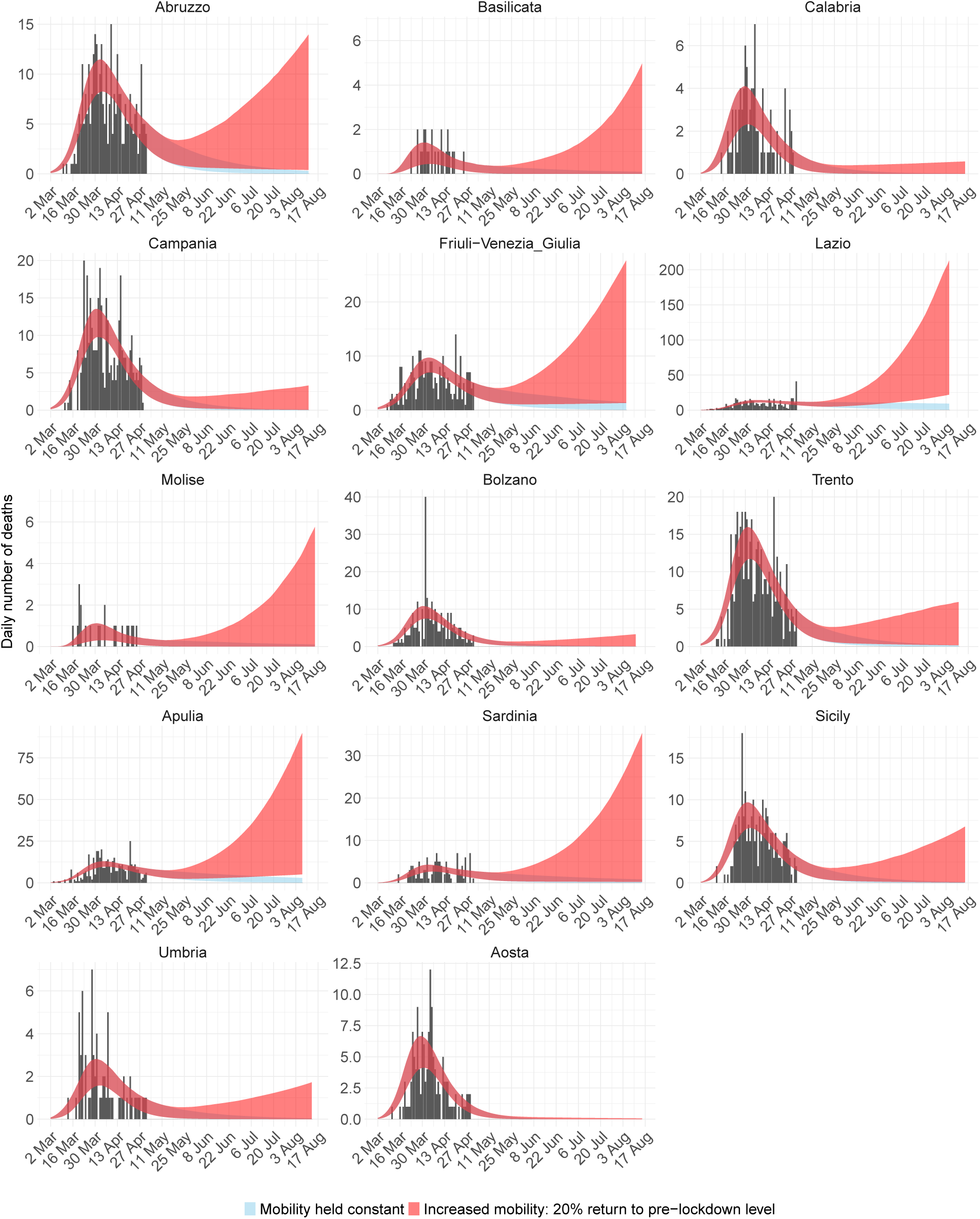
Deaths for the 14 Italian regions not included in the main text. Black bars are the data, red ribbon is the 95% credible interval forecast scenario were mobility stays the same, and blue is the 95% credible interval forecast scenario where mobility returns by 20% to pre-lockdown levels.

**Figure 15:**
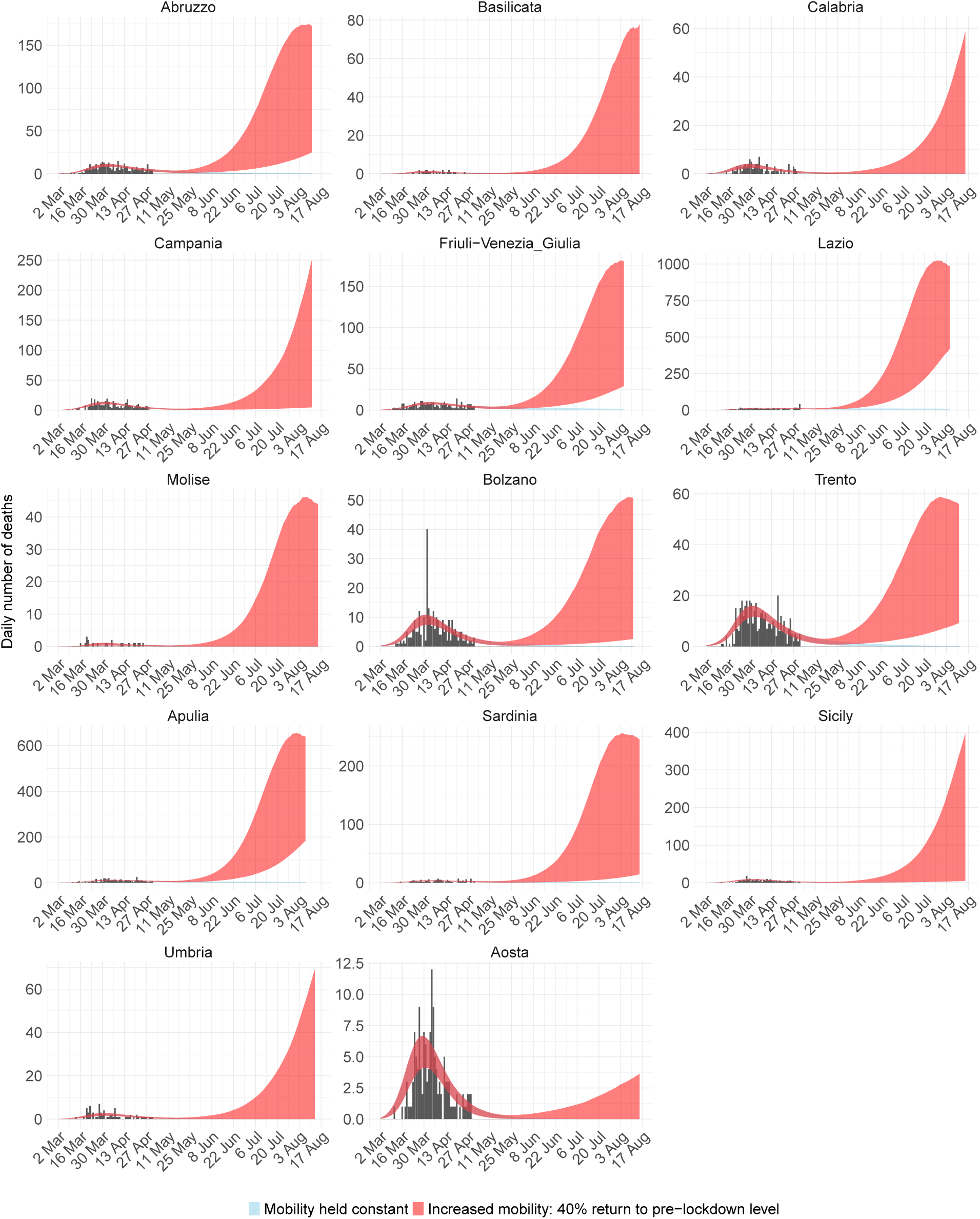
Deaths for the 14 Italian regions not included in the main text. Black bars are the data, red ribbon is the 95% credible interval forecast scenario were mobility stays the same, and blue is the 95% credible interval forecast scenario where mobility returns by 40% to pre-lockdown levels.

#### 6.3 Interventions

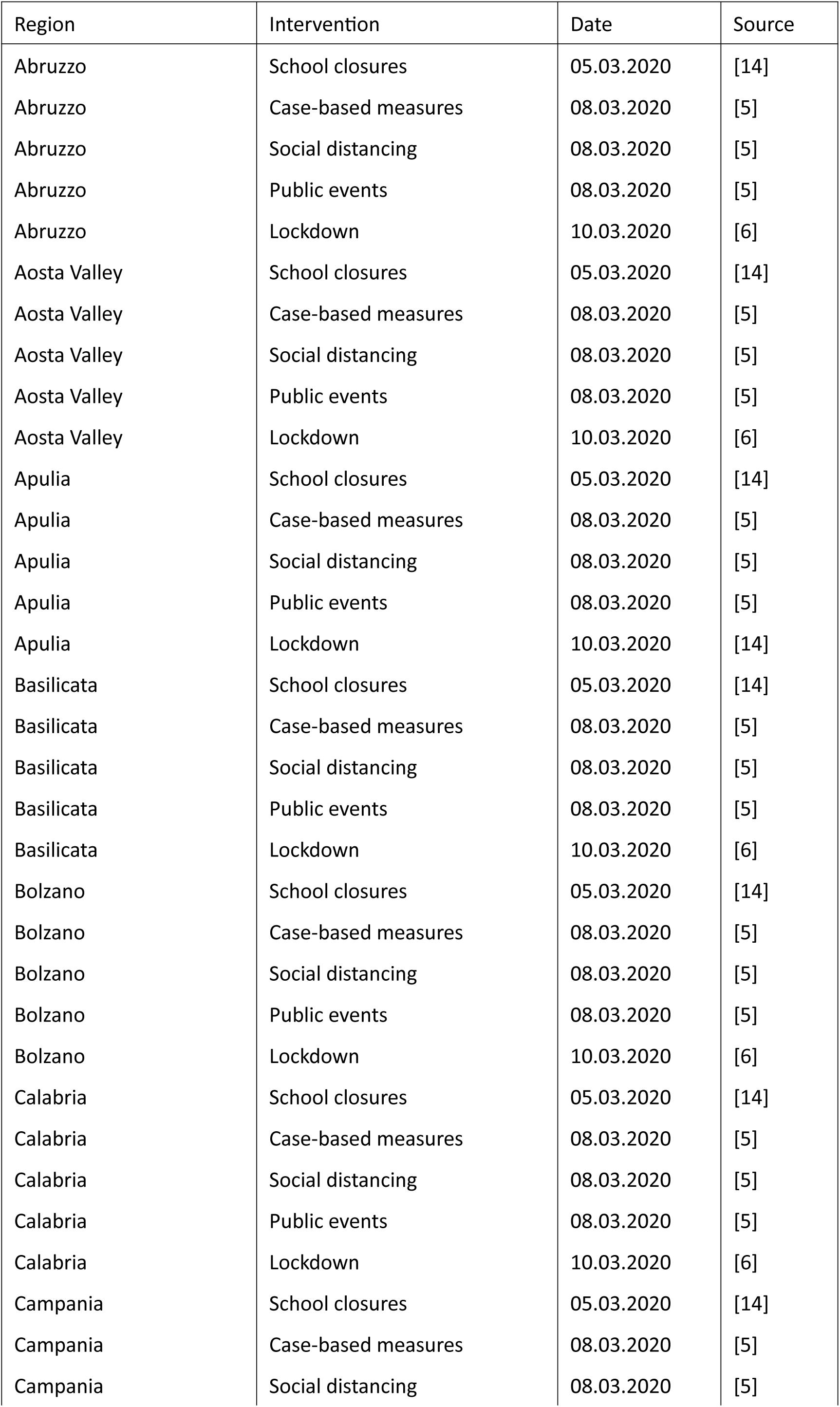

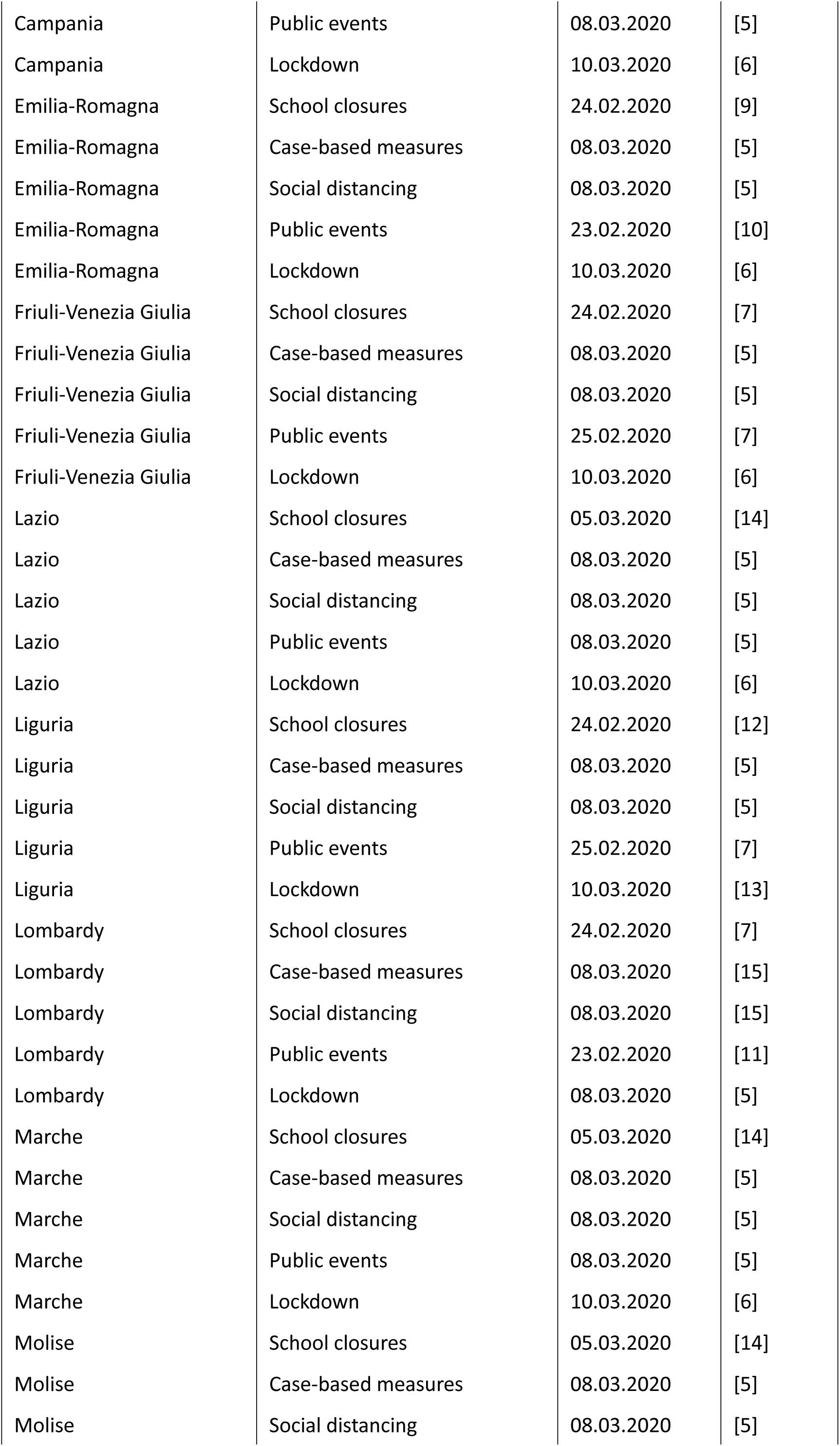

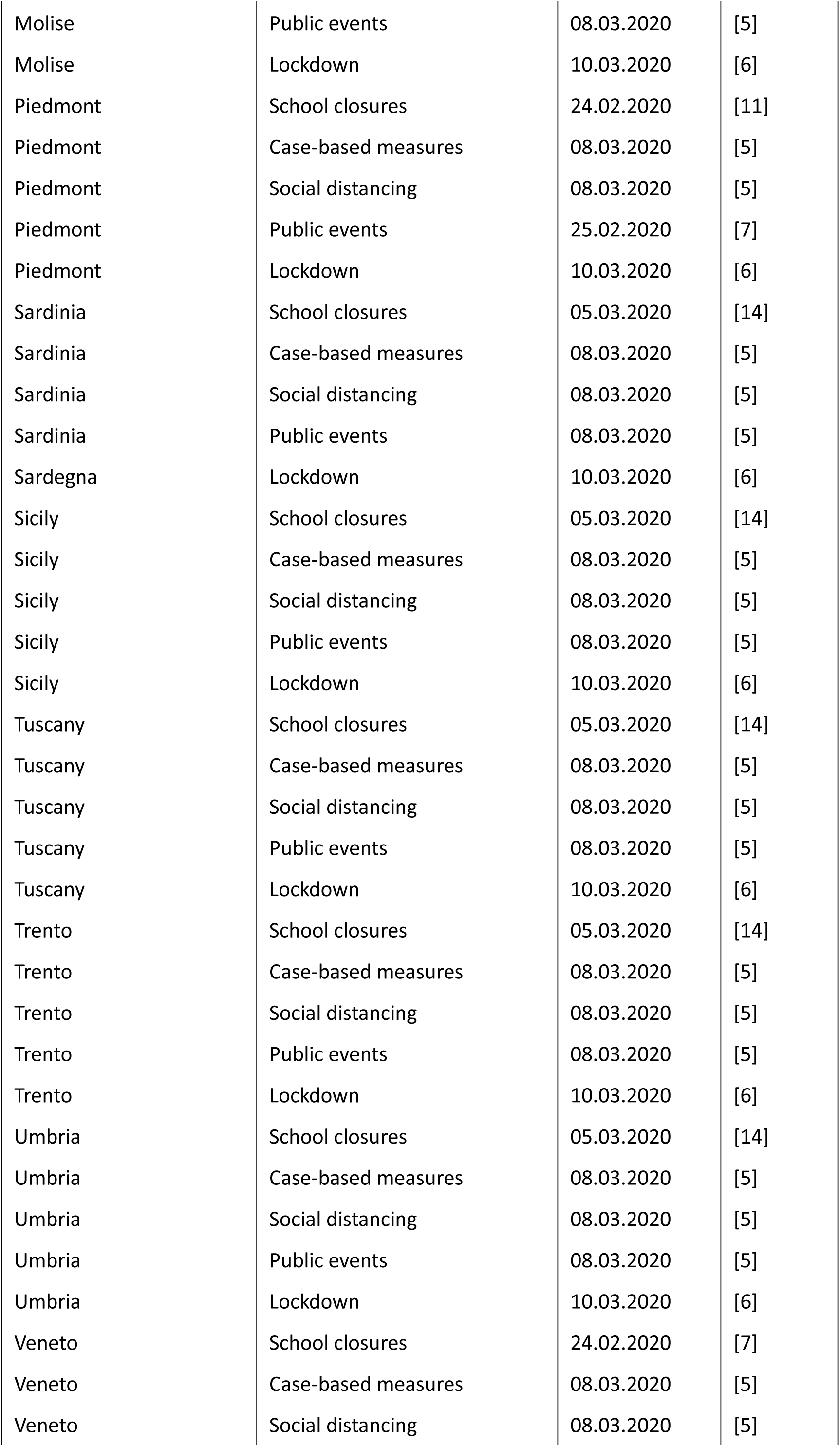

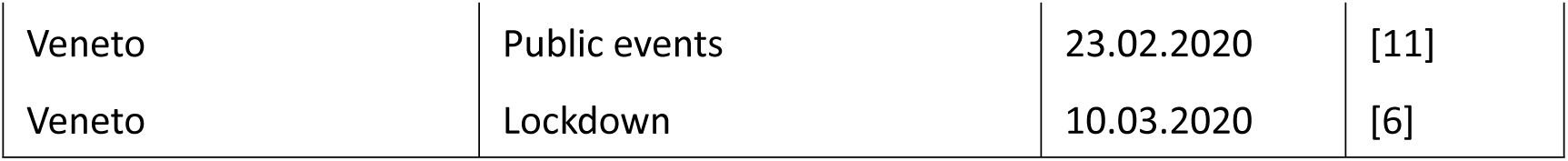

